# The Relationship Between Self-Determination and Burnout: Mental Health Outcomes in Medical Residents

**DOI:** 10.1101/2024.08.02.24311431

**Authors:** Hassan Mobarak, Chadia Haddad, Pascale Salameh, Evelyne Towair, Myriam El Khoury-Malhame, Rajaa Chatila

## Abstract

**Background:** Burnout is a pervasively increasing threat to personal and professional wellbeing and performance. It is yet understudied in relation to basic psychological needs (BPN), especially in at-risk population such as medical residents. This study intends to explore the differential relationship between various aspects of burnout including depersonalization (DP), emotional exhaustion (EE) and lack of personal achievement (PA) and subsets of BPN satisfaction or frustration namely autonomy, relatedness, and competence, with the framework of the Self-Determination Theory (SDT) in healthcare.

**Materials:** A total of 110 medical residents in various Lebanese hospitals were included. Demographics and standardized scales were used to measure basic psychological need satisfaction and frustration (BPNSFS), burnout (MBI), depression and anxiety (PHQ-4). Residents were also asked about subjective evaluation of academic training and level of impact by ongoing crises (COVID-19 pandemic, Beirut port explosion and financial breakdown).

**Results:** Result point to alarming prevalence of burnout and mental distress in our sample. It also indicates a differential correlation between gender, financial security and various subsets of burnout. It lastly points to association of DP with overall satisfaction scale (Beta=0.342, *p*=0.001) and PHQ-4 scores (Beta=-0.234, *p*=0.017), while feeling burdened to attend lectures and having been physically affected by the Beirut blast correlated with a sense of PA (Beta=0.332, *p*=0.010, Beta=0.187, *p*=0.041 respectively) and PHQ-4 (Beta=0.341, *p*=0.000), interacting with COVID-19 patients (Beta=0.168, *p*=0.020) and feeling protected in the working environment (Beta=-.231, *p*=0.002) showed a significant association with EE.

**Discussion:** Within the SDT framework, this study highlights the complex interplay between collective crises, subjective evaluations or work conditions and other demographics with aspects of burnout in medical residents. It mostly points to the need address this at an individual but also an institutional level to buffer distress in future healthcare providers.

## Introduction

Burnout is a state of physical, emotional and mental exhaustion (1–3). Since the emergence of the term in scientific literature, various theories have surfaced aiming to elucidate the origins for its development, including the Social Cognitive Theory, Social Exchange Theory, Organizational Theory, Structural Theory, Job Demands-Resources Theory, and Emotional Contagion Theory (4). More recently, the Self-Determination Theory (SDT), a theory for studying human motivation, has been popularly used as a promising theoretical framework to explain burnout (5–8). SDT is a meta-theory comprising five mini-theories, of which the basic psychological needs (BPNs) is the most implicated in the research on burnout (5,9,10). According to SDT, three inherent psychological needs are identified: autonomy, competence, and relatedness. The need for autonomy signifies an individual’s aspiration to self-regulate and exercise psychological freedom and choice in their professional activities. Competence pertains to a person’s desire to engage effectively with their medical responsibilities, experiencing mastery and confronting challenges. The need for relatedness represents the need for connection with colleagues and the cultivation of positive professional relationships (5). Individuals whose psychological needs are met tend to be more highly motivated. They actively participate in activities they find valuable, interesting, and enjoyable, indicating autonomous intrinsic motivation (11). Several studies showed that BPNs satisfaction is negatively correlated with burnout (8–10).

The high prevalence of burnout among medical professionals in general and medical residents in particular has been attributed to several factors (12–15). The demanding nature of residency training, which requires residents to acquire advanced medical knowledge and clinical skills all while also providing care for patients during clinical duties, contributes to heightened stress levels (16,17). This prolonged exposure to work stressors can lead to several negative outcomes on wellbeing such as burnout if not managed properly. The period of residency inherently encompasses substantial duties, necessitating prolonged working hours and call requirement obligations. This leads to sleep deprivation, loss of autonomy, and a sense of lack of control over one’s schedule (18–20).

Residents are often challenged with inadequate wages and imbalanced work/ home life. According to the Medscape Residents Lifestyle and Happiness Report, 2020, 27% of the residents in the United States of America indicated that they seldom or never had the opportunity to engage in a fulfilling social life; among these individuals, 68% reported failing relationships attributed to this factor (14).

Burnout among residents can have significant negative impact on both the individuals themselves and the healthcare system. Residents having burnout are at an elevated risk of various adverse outcomes, including unprofessional behaviors, strained relationships, stress, alcohol misuse, depression and even suicidal thoughts or actions (21–23). There are reports indicating that physicians experiencing high levels of distress and burnout are more likely to be implicated in medical errors (21).

The COVID-19 pandemic has created new challenges, especially for the healthcare sector (24). With trainees at the frontline of this pandemic, working hours and patient load have increased significantly. The surge in the number of cases, increased patient load and working hours have led to a further increase in the prevalence of burnout and stress among medical professionals, especially frontliners (25).

In Lebanon, around 70% of the population have experienced one or more of the following challenges: the direct impact of war, or its resulting calamities, the presence of an economic crisis, the unpredictability of the political situation and/ or to top it all, the trauma resulting from the catastrophic Beirut blast of 2020(26). All these conditions have made the working environment extremely challenging. In a study conducted to assess the prevalence of stress, burnout, and depression among residents, approximately 22% had battled depression during their years of postgraduate training, and 16% had moderate to severe anxiety. In addition, many trainees reported self-treating with psychotropic medications (27).

In addition to evaluating the prevalence of burnout among residents from various training programs in Lebanon, during this period of heightened stress and multiple adversities, we seek to examine the correlation between burnout levels and the three psychological needs depicted by the SDT. Additionally, we aim to assess the association between burnout and symptoms of anxiety and depression among this group of young physicians on one hand, and on the other, for the first time to the best of our knowledge, between burnout and subjective aspects of residents’ training and behavioral coping styles (Figure 1 and 2).

**Figure 1:**
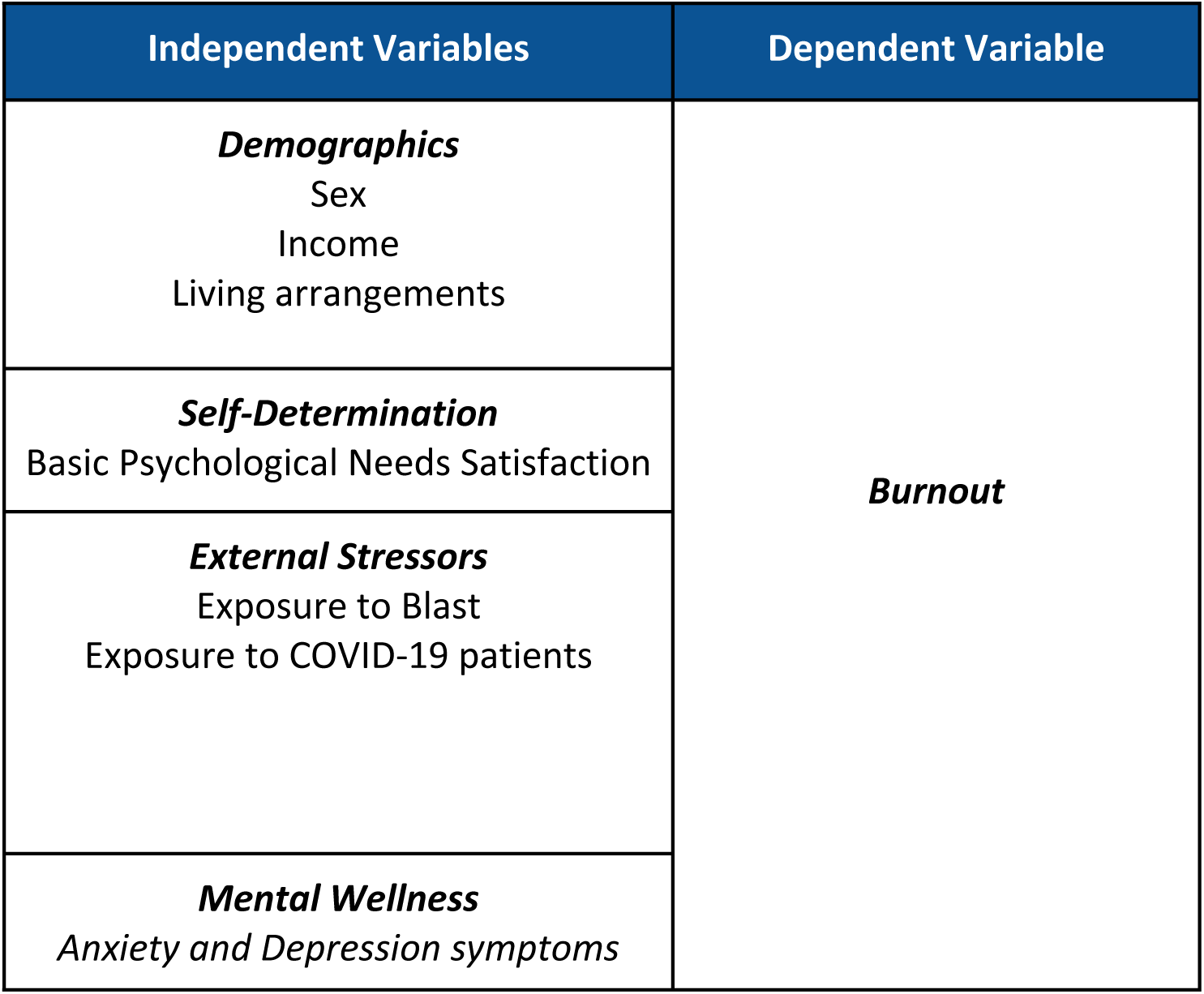
Studied variables.

**Figure 2:**
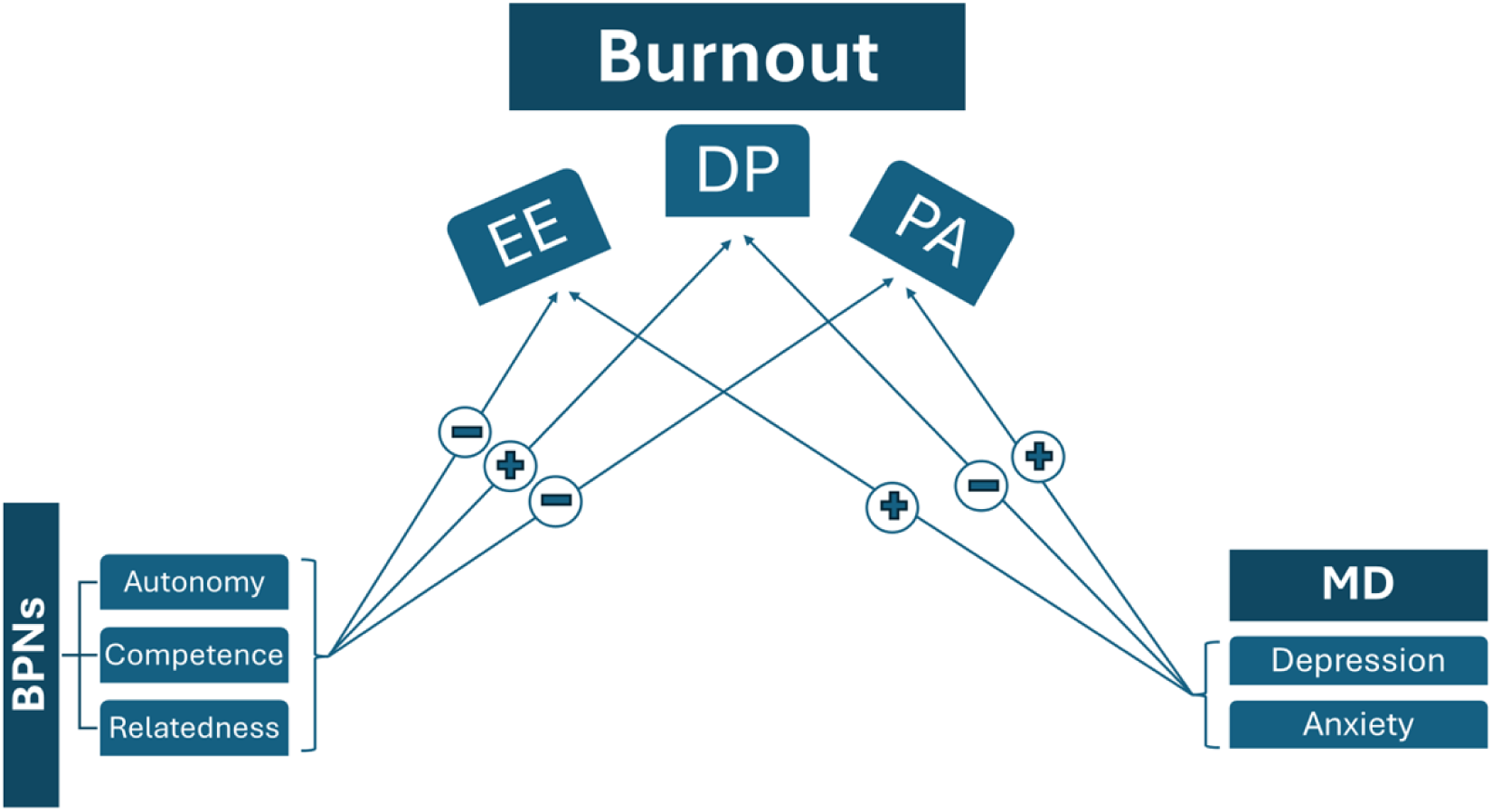
Suggested relationship between variables. BPNs: Basic Psychological Needs, EE: emotional Exhaustion, DP: depersonalization, PA: Personal achievement, MD: Mental Distress. (+): positively correlated, (-): negatively correlated.

## Methodology

### Study Design and Setting

After receiving approval from the Institutional Review Board at Lebanese American University (LAU.SOM.RC1.30/Dec/2020), we conducted a cross-sectional study of residents from multiple specialties in Lebanon. The recruitment process was done through social media platforms. The informed consent for participation in the study was obtained in electronically. All participants first provided a written consent by approving to participate and then were required to fill the anonymized online survey. Data was collected between September 2021 and March 2023.

### Study size

To estimate the minimal necessary sample size, the G*Power software Version 3.1.9.4 was used. In the absence of previous studies conducted in Lebanon related to the topic, we estimated the correlation between variables to be of moderate effect size (f2=0.15; squared multiple correlation r^2^=0.5), including 8 predictors per linear multiple regression model. Taking an alpha equal to 5% and a beta equal to 20%, a minimum sample size of n=109 is necessary to show the correlation between variables with a power equal to 0.8041.

### Participants

All residents who were directly or indirectly affected by the catastrophic events of 2020 (COVID-19 pandemic, Beirut blast, and economic crisis) were eligible for participation in this study. The research sample (N = 110) consisted of 51 females (45.5%) and 59 males (52.7%) after respondents with missing values were excluded (N=2). The age of the respondents ranged from 27 to 38 years (Mode=26, 23.2% of sample).

### Variables

A self-administered anonymous survey was developed. The survey consisted of three parts. Part one included general demographic information. Part two included questions on burnout, depression, anxiety, BPNs satisfaction, impact by COVID-19, economic crisis, and Beirut blast. Part three consisted of questions that explored individual behavioral coping, both adaptive and maladaptive, including for instance change in smoking and drinking habits, academic performance, and self-prescription of psychotropic medications. For under license surveys, due fees were paid to get the permission to administer the questionnaires in English.

### Burnout

There is no consensus on the methods used to evaluate burnout, nor on the cutoff levels (28–31). In this study, burnout was assessed using the Maslach Burnout Inventory-Human Services Survey for Medical Personnel (MBI-HSS[MP]) questionnaire, which is an adjusted version of Maslach Burnout Inventory (MBI) developed by Maslach and Jackson in 1986 (32). This questionnaire comprises 22 items that evaluate three dimensions: Emotional Exhaustion (EE), Depersonalization (DP), and Personal Achievement (PA). Participants rated their job-related feelings on a scale of 0 to 6, with 0 indicating never experiencing such feelings and 6 indicating experiencing them every day. Mean scores were computed for each subscale, indicating low, moderate, or high levels of EE, DP, and PA based on established scoring criteria. A positive indication of burnout was determined by moderate (17–26) or high (27+) scores on EE, moderate (7–12) or high (13+) scores on DP, or moderate (38–32) or low (0–31) scores on PA. Higher scores in EE and DP, along with lower scores in PA, are indicative of greater burnout.

### Basic Psychological Needs Satisfaction

Need satisfaction and frustration was measured using Basic Psychological Need Satisfaction and Frustration Scale (BPNSFS) by Chen et al (33). This scale has proven effective in various studies conducted among diverse populations, spanning both Western and non-Western demographics, including elementary school children and adults. These studies have utilized correlational methods, such as cross-sectional or diary studies, as well as experimental designs. They have explored the dynamics of needs at both a general level and within specific domains (e.g., physical education, relationships, training), as well as examining situational factors (34). The BPNSFS consists of 12 items reflecting need satisfaction and 12 items assessing need frustration. Each of the two parts consists of three dimensions: Autonomy (satisfaction and frustration), relatedness (satisfaction and frustration), and competence (satisfaction and frustration). Thus, the scale consists of 24 items representing 6 dimensions (4 items each). Example items are “I feel a sense of choice and freedom in the things I undertake” (autonomy satisfaction), “I am confident that I can do things well” (competence satisfaction), and “I feel connected to people who care about me and who I also care about” (relatedness satisfaction). The answers or statements refer to the current life situation and are given on a scale from 1 (completely untrue) to 5 (completely true). The higher the result on a subscale is, the stronger is the satisfaction or frustration of the need. The BPNSFS does not have standardized cutoff values. The scale is typically used to assess relative levels of need satisfaction and frustration rather than to classify individuals into categories(35).

### Anxiety and Depression

Anxiety and depression symptoms were assessed using the Patient Health Questionnaire-4 (PHQ-4). This tool comprises self-reported questionnaires: the 2-item Patient Health Questionnaire (PHQ-2) and the 2-item Generalized Anxiety Disorder screening tool (GAD-2) (36). The PHQ-2 includes the two main diagnostic criteria for depressive disorders according to the DSM-V, while the GAD-2 covers the main criteria for Generalized Anxiety Disorder. Several studies have shown that these tools are reliable for screening (36–40). The combined results of these two-item measures constitute the PHQ-4. For both the PHQ-2 and GAD-2, a sum score of three or higher was recommended as the cut-off point for probable depression or anxiety, respectively, based on the study by Kroenke et al. in 2009 (36).

### Statistical Methods

All analyses were carried out using the Statistical Package for Social Sciences (version 24.0). For descriptive analysis, frequency and percentage were used for categorical variables, and mean and standard deviation for quantitative variables. The distribution of continuous variables was considered normal using visual inspection of the histogram, while the skewness and kurtosis were lower than 1.

For the bivariate analysis of continuous variables, the Student’s T-test was used to compare the means between 2 groups and ANOVA to compare between three groups or more, after checking for homogeneity of variances using Levene’s test. In case the variances are not homogenous, the corrected T-test and the Kruskal-Wallis test were used, respectively. After ANOVA and Kruskal-Wallis significant testing, post hoc analyses were conducted using Bonferroni adjustment. A Spearman correlation coefficient was used between continuous variables, and a gamma coefficient to assess the association between ordinal variables. Categorical variables association was assessed using the Chi-square test.

As for the multivariable analysis, multiple linear regressions using the General Linear Model process were conducted to assess the correlates of dependent variables in the whole sample, after checking the residues’ normality, the linearity of the relationship, the absence of multicollinearity and the homoscedasticity assumptions; a stepwise method was used to reach the most parsimonious model. The beta coefficient, its 95% Confidence Interval and the p = were reported. Independent variables introduced in the models will be those that have a p = lower than 0.1 in the bivariate analysis, considering the maximum number allowed of variables to be included given the sample size: sociodemographic, and other independent variables were added as appropriate.

## Results

### Demographic Data

A total of 110 residents/trainees participated in the study. The mean (±SD) age was 28.01 years (±9.21). Almost half the participants were males (52.7%), single (65.2%) and 47.3% were living with their parents. As for the household income, 33% had an income > 10 million Lebanese Pounds, 10.7% did not provide an answer regarding their monthly income. The majority of participants were in postgraduate year 3 and below (72.7%) and were rotating in hospitals within Beirut at the time of the Beirut blast. (Table 1)

**Table 1:** Sociodemographic Characteristics.

| **Table 1: Sociodemographic Characteristics** |  |
| --- | --- |
|  | **Frequency (%)** |
| **Gender** |  |
| Female | 51 (45.5%) |
| Male | 59 (52.7%) |
| **Marital status** |  |
| In a stable relationship | 23 (20.5%) |
| Married | 14 (12.5%) |
| Single | 73 (65.2%) |
| **Living status** |  |
| Alone | 30 (26.8%) |
| With a Flat mate | 14 (12.5%) |
| With a partner | 13 (11.6%) |
| With parents | 53 (47.3%) |
| **Average household Income** |  |
| < 2.5 million LBP/month | 6 (5.5%) |
| Between 2.5-5 million LBP/ month | 24 (21.8%) |
| Between 5-10 million LBP/month | 31 (28.2%) |
| >10 million LBP/month | 37 (33.6%) |
| Prefer not to answer | 12 (10.9%) |
| **Postgraduate year** |  |
| PGY 1 | 34 (30.9%) |
| PGY 2 | 20 (18.2%) |
| PGY 3 | 29 (26.4%) |
| PGY 4 | 14 (12.7%) |
| PGY 5 | 6 (5.5%) |
| PGY 6 | 5 (4.5%) |
| Other | 2 (1.8%) |
| **Rotation in Beirut During Beirut Blast** |  |
| Yes | 80 (72.7%) |
| No | 30 (27.3%) |
|  | **Mean ± SD** |
| **Age** | 28.10 ± 9.21 |

LBP: Lebanese pound; PGY: postgraduate year; SD: standard deviation

### BPNs

The results of the Basic Psychological Need Satisfaction and Frustration Scale (BPNSFS) indicate varied levels of satisfaction and frustration across different dimensions (Table 2). BPS for the needs of autonomy, relatedness, and competence had a mean of 13.67 (± 3.13), 14.49 (±3.42), and 15.00 (±3.55) respectively where 20 is the maximum grade of each of them. While the frustration of the same needs had a mean score of 13.02 (±3.80), 9.53 (±4.22), and 11.98 (±2.82). The overall satisfaction which is the satisfaction of all 3 needs combined was 42.78 (±9.05) where maximum value is 60.

**Table 2.**
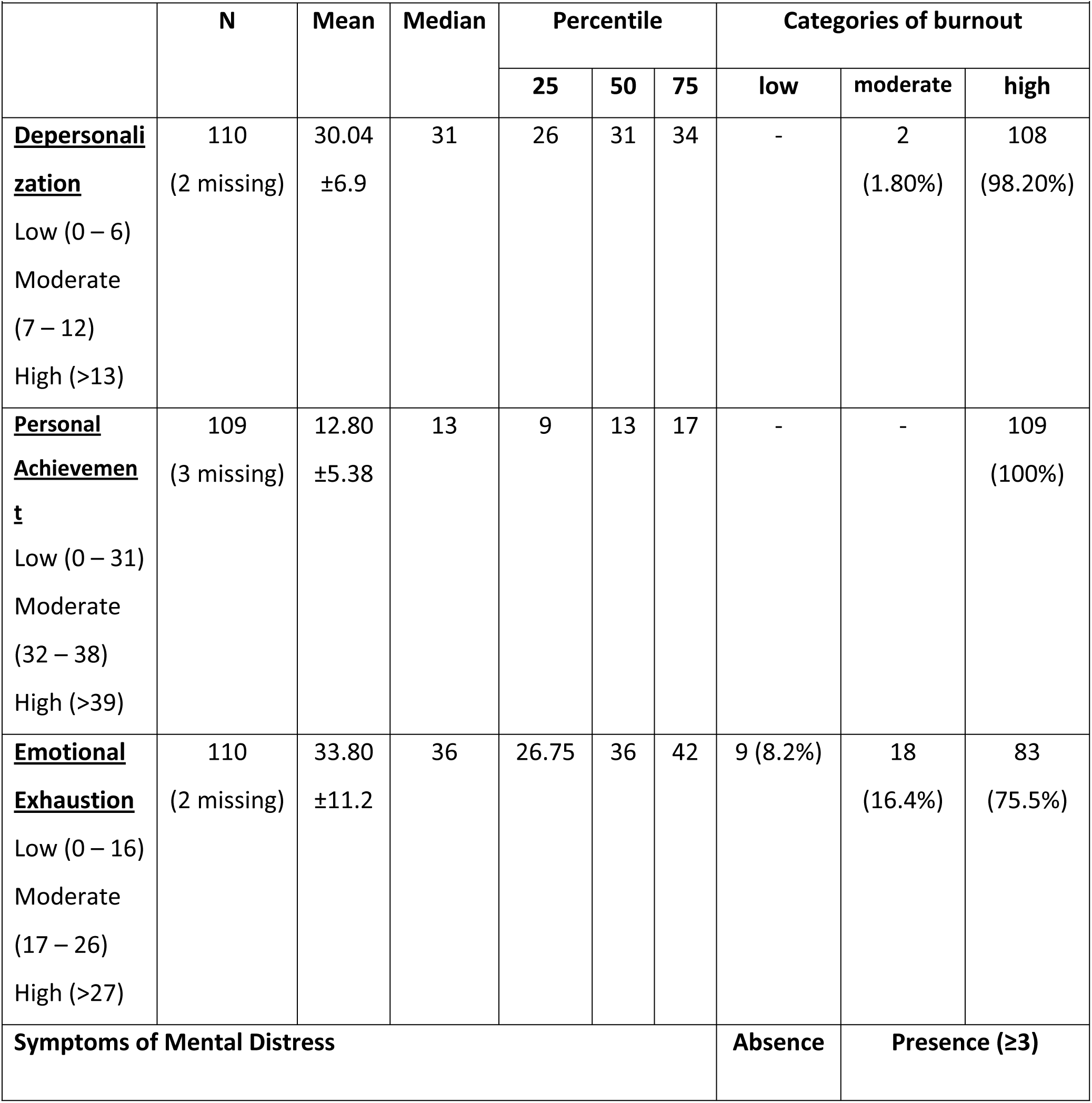
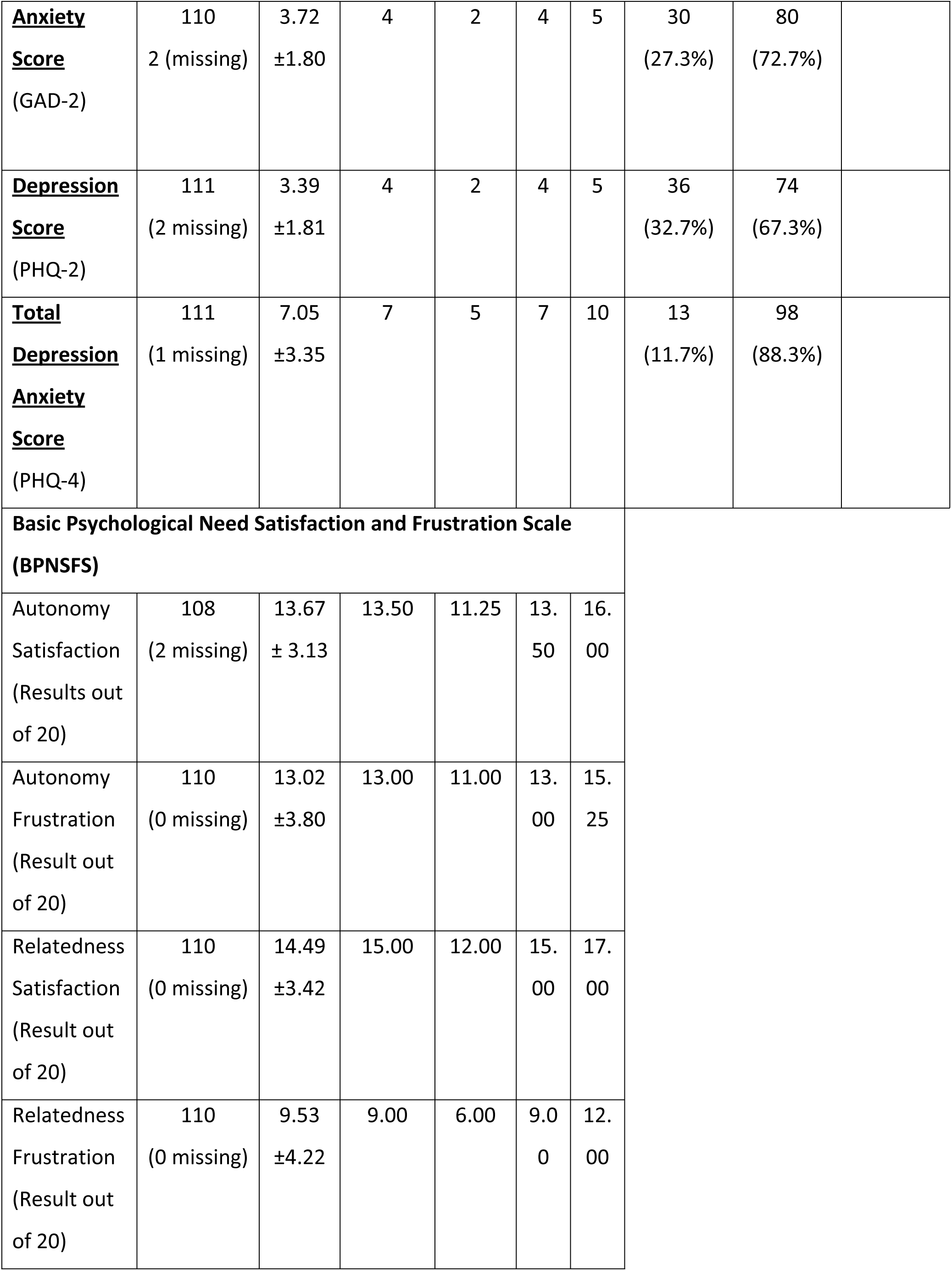
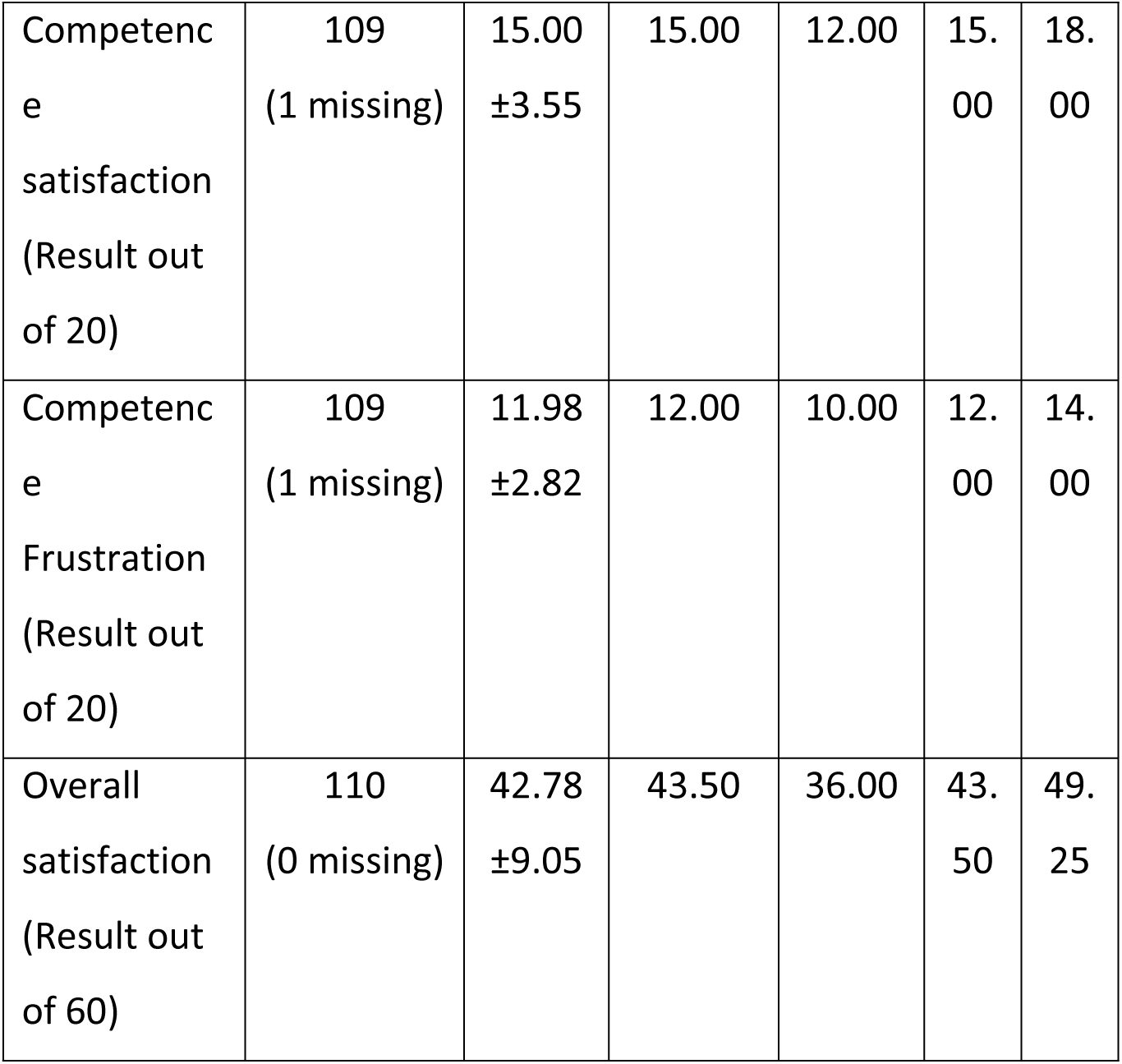
Prevalence and Severity of the 3 Dimensions of the Burnout.

| **Table 2. Prevalence and Severity of the 3 Dimensions of the Burnout** |  |  |  |  |  |  |  |  |  |
| --- | --- | --- | --- | --- | --- | --- | --- | --- | --- |
|  | N | Mean | Median | Percentile |  |  | Categories of burnout |  |  |
|  |  |  |  | 25 | 50 | 75 | low | moderate | high |
| **<u>Depersonalization</u>**<br>Low (0 – 6)<br>Moderate (7 – 12)<br>High (>13) | 110<br>(2 missing) | 30.04<br>±6.9 | 31 | 26 | 31 | 34 | - | 2<br>(1.80%) | 108<br>(98.20%) |
| **<u>Personal Achievement</u>**<br>Low (0 – 31)<br>Moderate (32 – 38)<br>High (>39) | 109<br>(3 missing) | 12.80<br>±5.38 | 13 | 9 | 13 | 17 | - | - | 109<br>(100%) |
| **<u>Emotional Exhaustion</u>**<br>Low (0 – 16)<br>Moderate (17 – 26)<br>High (>27) | 110<br>(2 missing) | 33.80<br>±11.2 | 36 | 26.75 | 36 | 42 | 9 (8.2%) | 18<br>(16.4%) | 83<br>(75.5%) |
| **Symptoms of Mental Distress** |  |  |  |  |  |  | **Absence** | **Presence (≥3)** |  |
| **<u>Anxiety</u>**<br>**<u>Score</u>**<br>(GAD-2) | 110<br>2 (missing) | 3.72<br>±1.80 | 4 | 2 | 4 | 5 | 30<br>(27.3%) | 80<br>(72.7%) |  |
| **<u>Depression</u>**<br>**<u>Score</u>**<br>(PHQ-2) | 111<br>(2 missing) | 3.39<br>±1.81 | 4 | 2 | 4 | 5 | 36<br>(32.7%) | 74<br>(67.3%) |  |
| **<u>Total</u>**<br>**<u>Depression</u>**<br>**<u>Anxiety</u>**<br>**<u>Score</u>**<br>(PHQ-4) | 111<br>(1 missing) | 7.05<br>±3.35 | 7 | 5 | 7 | 10 | 13<br>(11.7%) | 98<br>(88.3%) |  |
| **Basic Psychological Need Satisfaction and Frustration Scale**<br>**(BPNSFS)** |  |  |  |  |  |  |  |  |  |
| Autonomy<br>Satisfaction<br>(Results out<br>of 20) | 108<br>(2 missing) | 13.67<br>± 3.13 | 13.50 | 11.25 | 13.<br>50 | 16.<br>00 |  |  |  |
| Autonomy<br>Frustration<br>(Result out<br>of 20) | 110<br>(0 missing) | 13.02<br>±3.80 | 13.00 | 11.00 | 13.<br>00 | 15.<br>25 |  |  |  |
| Relatedness<br>Satisfaction<br>(Result out<br>of 20) | 110<br>(0 missing) | 14.49<br>±3.42 | 15.00 | 12.00 | 15.<br>00 | 17.<br>00 |  |  |  |
| Relatedness<br>Frustration<br>(Result out<br>of 20) | 110<br>(0 missing) | 9.53<br>±4.22 | 9.00 | 6.00 | 9.0<br>0 | 12.<br>00 |  |  |  |

|  |  |  |  |  |  |  |
| --- | --- | --- | --- | --- | --- | --- |
| Competence satisfaction<br>(Result out of 20) | 109<br>(1 missing) | 15.00<br>$\pm 3.55$ | 15.00 | 12.00 | 15.00 | 18.00 |
| Competence Frustration<br>(Result out of 20) | 109<br>(1 missing) | 11.98<br>$\pm 2.82$ | 12.00 | 10.00 | 12.00 | 14.00 |
| Overall satisfaction<br>(Result out of 60) | 110<br>(0 missing) | 42.78<br>$\pm 9.05$ | 43.50 | 36.00 | 43.50 | 49.25 |

### Burnout Dimensions

All three dimensions of burnout were highly prevalent in the study population. 98.20% exhibiting high levels of DP on the MBI scale with a mean DP score being 30.04 (± 6.9). All participants had a low sense of PA with a mean of 12.80 (±5.38). For EE, 91.9% of participants had moderate or high levels with a mean score of 33.80 (±11.2) while only 8.2% reported low levels of EE (Table 2).

### Anxiety and Depression

The prevalence of generalized anxiety, depression symptoms and total depression/anxiety as measured by the GAD-2 and PHQ-2 and PHQ-4 were 72.7%, 67.3% and 88% respectively, with mean values of 3.72 (±1.80), 3.39 (±1.81) and 7.05 (±3.35) respectively.

### Bivariate analysis

#### Demographic factors and burnout dimensions

There were no notable gender disparities in burnout except for a slight variation in DP levels, with males experiencing higher levels (with a mean of 31.30 (±7.17) compared to females 28.56 (±6.34), (p= 0.038) (Table 3). Living status played a role in the DP dimension of burnout, where those who live with a partner or live alone had the highest means of DP (34.30 (±6.40) and 30.83 (±4.32) respectively) (*p*=0.025). On the other hand, participants with an average income of < 2.5 Million LBP/month had the highest level of EE of 42.16 (±11.08, *p*=0.031). Marital status, year of training, and being in Beirut during the blast had no significant impact on the levels of burnout.

**Table 3:** Bivariate Analysis with Burnout Dimensions as the Dependent Variables.

|  | DP | p-value | PA | p-value | EE | p-value |
| --- | --- | --- | --- | --- | --- | --- |
|  | Mean ± SD |  | Mean ± SD |  | Mean ± SD |  |
| **Gender** |  |  |  |  |  |  |
| Female | 28.56±6.34 | **0.038** | 12.24±4.35 | 0.309 | 35.70±9.94 | 0.099 |
| Male | 31.30±7.17 |  | 13.27±6.12 |  | 32.18±12.19 |  |
| **Marital status** |  |  |  |  |  |  |
| In a stable relationship | 29.34±5.61 | 0.168 | 12.69±5.30 | 0.637 | 35.73±9.97 | 0.633 |
| Married | 33.28±7.36 |  | 11.57±5.44 |  | 32.50±10.68 |  |
| Single | 29.63±7.10 |  | 13.06±5.44 |  | 33.46±11.85 |  |
| **Living Status** |  |  |  |  |  |  |
| Alone | 30.83±4.32 | **0.025** | 12.63±4.73 | 0.362 | 33.53±13.39 | 0.821 |
| With a flat mate | 26.64±8.50 |  | 12.92±6.29 |  | 34.00±11.46 |  |
| With a partner | 34.30±6.40 |  | 10.46±5.30 |  | 31.23±10.94 |  |
| With parents | 29.43±7.32 |  | 13.44±5.49 |  | 34.56±10.22 |  |
| **Average Household Income** |  |  |  |  |  |  |
| < 2.5 million LBP/month | 25.00±8.94 | 0.171 | 17.00±7.42 | 0.255 | 42.16±11.08 | **0.031** |
| 2.5-10 million LBP/ month | 29.96±6.43 |  | 12.50±4.76 |  | 35.54±9.69 |  |
| >10 million LBP/month | 31.37±6.33 |  | 12.81±5.56 |  | 30.05±11.70 |  |
| Prefer not to answer | 28.75±8.93 |  | 12.00±6.22 |  | 33.33±14.06 |  |
| **Postgraduate Year** |  |  |  |  |  |  |
| PGY 1 | 28.97±7.49 | 0.640 | 13.17±5.58 | 0.388 | 31.55±10.85 | 0.572 |
| PGY 2 | 29.65±8.40 |  | 14.30±5.39 |  | 35.30±7.98 |  |
| PGY 3 | 30.51±4.88 |  | 12.32±5.20 |  | 34.82±13.10 |  |
| ≥PGY 4 | 31.14±6.95 |  | 11.70±5.31 |  | 34.48±11.99 |  |
| Rotation in Beirut During Beirut Blast |  |  |  |  |  |  |
| Yes | 30.63±6.65 | 0.137 | 12.75±5.82 | 0.888 | 33.65±11.20 | 0.800 |
| No | 28.43±7.43 |  | 12.90±4.09 |  | 34.26±11.71 |  |
|  | DP<br>Correlation<br>coefficient | p-value | PA<br>Correlatio<br>n<br>coefficient | p-value | EE<br>Correlatio<br>n<br>coefficient | p-value |
| Age | 0.011 | 0.908 | -0.107 | 0.268 | -0.113 | 0.240 |
| Autonomy Satisfaction | 0.434 | <0.001 | -0.217 | 0.025 | -0.397 | <0.001 |
| Autonomy Frustration | -0.217 | 0.234 | 0.466 | <0.001 | 0.710 | <0.001 |
| Relatedness Satisfaction | 0.377 | <0.001 | -0.190 | 0.048 | -0.294 | 0.002 |
| Relatedness Frustration | -0.281 | 0.003 | 0.414 | <0.001 | 0.262 | 0.006 |
| Competence satisfaction | 0.508 | <0.001 | -0.209 | 0.030 | -0.270 | 0.005 |
| Competence Frustration | -0.161 | 0.094 | 0.379 | <0.001 | 0.396 | <0.001 |
| Overall satisfaction | 0.497 | <0.001 | -0.304 | 0.001 | -0.407 | <0.001 |
| Anxiety Score (GAD-2) | -0.257 | 0.007 | 0.149 | 0.122 | 0.480 | <0.001 |
| Depression Score (PHQ-2) | -0.379 | <0.001 | 0.362 | <0.001 | 0.638 | <0.001 |
| Total Depression Anxiety Score (PHQ-4) | -0.354 | <0.001 | 0.297 | 0.002 | 0.622 | <0.001 |
| Level of interaction with patients who were COVID-19 positive | 0.098 | 0.308 | 0.231 | **0.016** | 0.211 | **0.027** |
| Having access to PPE for protection | -0.018 | 0.855 | -0.169 | 0.079 | -0.201 | **0.035** |
| Being infected by COVID 19 | -0.109 | 0.269 | -0.015 | 0.883 | 0.066 | 0.501 |
| Family being infected by COVID 19 | 0.086 | 0.379 | -0.008 | 0.932 | 0.030 | 0.756 |
| Physically affected by Beirut Blast | 0.008 | 0.935 | 0.269 | **0.005** | 0.145 | 0.131 |
| Emotionally affected by Beirut Blast | -0.118 | 0.219 | 0.205 | **0.032** | 0.294 | **0.002** |
| Financially affected by Beirut Blast | -0.010 | 0.917 | 0.173 | 0.073 | 0.182 | 0.058 |
| Family affected by Beirut Blast | 0.050 | 0.604 | 0.027 | 0.780 | 0.078 | 0.418 |
| Being personally affected by economic crisis | 0.165 | 0.085 | 0.055 | 0.568 | 0.237 | **0.013** |
| Family's financial support being adequate | 0.040 | 0.681 | -0.142 | 0.141 | -0.55 | 0.105 |
| Overall income able to cover essential daily needs | 0.036 | 0.711 | -0.128 | 0.184 | -0.327 | **<0.001** |
| Feeling adequately prepared for In-training exam | 0.206 | **0.031** | -0.263 | **0.0006** | -0.435 | **<0.001** |
| Feeling burdened by attending lectures | -0.089 | 0.360 | 0.354 | **<0.001** | 0.380 | **<0.001** |
| The adherence to attending lectures affected | 0.104 | 0.283 | 0.229 | **0.016** | 0.327 | **0.001** |
| Enthusiasm to offer clinical care affected | -0.133 | 0.166 | 0.171 | 0.076 | 0.444 | **<0.001** |
| Increase frequency of smoking | 0.026 | 0.784 | 0.131 | 0.175 | 0.023 | 0.813 |
| Increase alcohol consumption | -0.019 | 0.841 | 0.054 | 0.577 | -0.008 | 0.932 |
| Resorted to self-medication | -0.090 | 0.348 | 0.217 | **0.023** | 0.228 | **0.016** |

#### Psychological needs satisfaction, specific residency-related stressors and burnout dimensions

There was a significant correlation between satisfaction of the 3 psychological needs and burnout dimensions (Table 3), with the correlation being positive with DP and negative with EE and PA.

Mental distress symptoms (PHQ-4) also negatively correlated with DP (r=-0.354, *p*<0.001), while positively correlating with PA (r=0.297, *p*=0.002) and EE (r=0.622, *p*<0.001).

There was a positive correlation between both the level of interaction with COVID-19 patients and feeling burdened by attending lectures with the sense of PA (r=0.231, *p*=0.016 and r=0.354, *p*<0.001 respectively). On the other hand, residents feeling adequately prepared for their in-training exam negatively correlated with PA (r=-0.263, *p*=0.0006) and had lower levels of EE (r=-0.435, *p*<0.001). Residents experienced more EE if they were burdened by attending lectures (r=0.380, *p*<0.001) and if their enthusiasm to offer clinical care was affected (r=0.444, *p*<0.001). Lastly, those adequately prepared for exams had higher DP (r=0.206, *p*=0.031).

### Multivariate Analysis

To explore factors influencing burnout dimensions and differentiating predictors and protective factors, a linear regression multivariate analysis was undertaken with the dependent variables being the three dimensions of burnout.

The Overall Satisfaction Scale demonstrated a statistically significant positive association with DP (Beta=0.342, *p*=0.001) (table 4). Conversely, thePHQ-4 scores exhibited a significant negative association with DP (Beta=-0.234, *p*=0.017).

**Table 4:** Multivariable Linear Regression.

| **Table 4: Multivariable Linear Regression** |  |  |  |  |  |
| --- | --- | --- | --- | --- | --- |
| **Model 1: Depersonalization as the Dependent Variable** |  |  |  |  |  |
|  | **Unstandardized Beta** | **Standardized Beta** | **p-value** | **Confidence interval** |  |
|  |  |  |  | **Lower Bound** | **Upper Bound** |
| Gender (Female vs Male*) | -.529 | -.038 | .660 | -2.912 | 1.853 |
| Living status (with a Flat-mate vs alone*) | -2.306 | -.112 | .254 | -6.293 | 1.681 |
| Living status (With a Partner vs alone*) | 1.879 | .088 | .355 | -2.129 | 5.886 |
| Living Status (With Parents vs alone*) | -.191 | -.014 | .892 | -2.973 | 2.591 |
| **Overall satisfaction scale** | **.261** | **.342** | **.001** | **.114** | **.407** |
| **Total depression anxiety (PHQ-4)** | **-.489** | **-.234** | **.017** | **-.889** | **-.089** |
| Affected by the financial crisis? | .244 | .071 | .418 | -.351 | .838 |
| Felt you can adequately prepare for your in-training exam? | .065 | .026 | .775 | -.387 | .517 |
| **Model 2: Personal Achievement as the Dependent Variable** |  |  |  |  |  |
| **Overall satisfaction scale** | -.065 | -.112 | .248 | -.177 | .046 |
| Total depression anxiety (PHQ-4) | .161 | .101 | .333 | -.168 | .490 |
| Interact with patients who are COVID positive? | .280 | .144 | .109 | -.063 | .624 |
| Protected in your working environment (PPE availability, vaccinations access, support)? | -.314 | -.115 | .189 | -.786 | .158 |
| **Physically affected by the Beirut blast** | **.608** | **.187** | **.041** | **.026** | **1.189** |
| Emotionally affected by the Beirut blast ? | .192 | .088 | .341 | -.206 | .589 |
| Financially affected by the Beirut blast? | -.056 | -.034 | .719 | -.365 | .253 |
| Felt you can adequately prepare for your in-training exam? | -.232 | -.121 | .195 | -.585 | .121 |
| **Burden you to attend your required lectures?** | **.602** | **.332** | **.010** | **.147** | **1.058** |
| Attendance of lectures was affected? | -.188 | -.104 | .406 | -.637 | .260 |
| Enthusiasm to offer clinical care was affected? | .057 | .031 | .730 | -.272 | .387 |
| Resorted to self-medicating? | .434 | .123 | .168 | -.187 | 1.055 |
| **Model 3: Emotional Exhaustion as the Dependent Variable** |  |  |  |  |  |
| Gender (Female vs Male*) | -.652 | -.029 | .696 | -3.957 | 2.654 |
| Income cat (Between 2.5M - 10M LBP / Month vs <2.5M*) | -.218 | -.004 | .958 | -8.461 | 8.026 |
| Income cat (=> 10M / Month vs <2.5M*) | -1.284 | -.057 | .627 | -6.520 | 3.952 |
| Income cat (prefer not to answer vs <2.5M*) | -.451 | -.019 | .870 | -5.925 | 5.022 |
| **Overall satisfaction scale** | -.158 | -.127 | .108 | -.350 | .035 |
| **Total depression anxiety (PHQ-4)** | **1.155** | **.341** | **.000** | **.593** | **1.717** |
| **Interact with patients who are COVID positive?** | **.695** | **.168** | **.020** | **.111** | **1.279** |
| **Protected in your working environment** | **-1.337** | **-.231** | **.002** | **-2.189** | **-.486** |
| Emotionally affected by the Beirut blast? | .562 | .121 | .109 | -.128 | 1.252 |
| Financially affected by the Beirut blast? | -.139 | -.040 | .584 | -.642 | .364 |
| Affected by the financial crisis? | .181 | .032 | .671 | -.661 | 1.023 |
| Overall income covers your essential daily needs? | -.376 | -.089 | .233 | -.999 | .247 |
| Felt you can adequately prepare for your in-training exam? | -.447 | -.109 | .147 | -1.054 | .160 |
| How much did it burden you to attend your required lectures? | .184 | .048 | .634 | -.582 | .951 |
| Feel that your attendance of lectures was affected? | .252 | .066 | .506 | -.497 | 1.000 |
| **Enthusiasm to offer clinical care was affected?** | **1.017** | **.261** | **.001** | **.446** | **1.588** |
| Resorted to self-medicating? | .705 | .094 | .180 | -.333 | 1.743 |

Feeling burdened to attend lectures and having been physically affected by the Beirut blast correlated with a sense of PA (Beta=0.332, *p*=0.010, Beta=0.187, *p*=0.041 respectively) (Table 4).

Total Depression Anxiety symptoms (PHQ-4) showed a significant positive association with EE (Beta=0.341, *p*=0.000), similarly did interacting with COVID-19 patients (Beta=0.168, *p*=0.020). In contrast, feeling protected in the working environment showed a significant negative association with EE (Beta=-.231, *p*=0.002) (Table 4).

## Discussion

To our knowledge, this is the first study to examine the relationship between self-determination, measured by the satisfaction of the three psychological needs, and burnout in medical residents in times of accumulating adversities. Our findings indicate a significant relationship between these variables, such as when the basic psychological needs are met, there is a reduction in EE, the main dimension of burnout that correlates with anxiety and depression symptoms.

SDT posits that the satisfaction of BPNs—autonomy, competence, and relatedness—is essential for psychological wellbeing and optimal functioning. However, the interplay between BPNs and burnout, especially within the context of medical residency where coping mechanisms and the complex nature of work demands can affect this dynamic, remains underexplored. In our study, we found that when the BPNs are met, residents are less likely to experience EE yet more likely to experience DP. In fact, healthcare workers are primarily driven by a mission to help and save and/or rescue others. It is possible that in high-risk situations where their personal safety is compromised, DP may emerge as a protective mechanism. This can lead to extreme dissociation, where healthcare workers do their jobs correctly but disconnect from reality, becoming impersonal or callous to cope with the distress of handling many patients or dealing with multiple stressors simultaneously.

While depersonalization might initially provide some relief from the overwhelming emotional demands of work, it is not a sustainable solution. Depersonalization can lead to a lack of engagement and a decrease in the quality of care or work performance, which can create a cycle of increasing stress and emotional exhaustion (41).

Albeit not surprising given the overall context, our study shows alarming rates of burnout among medical residents in our sample with almost all residents experiencing DP, EE and low PA. A recent study assessing burnout among postgraduate medical trainees in Lebanon similarly found that prevalence rates of high burnout are 37.2% for disengagement and 51.1% for exhaustion using a different scale at the early stages of the pandemic (42). Even prior to the COVID-19 viral spread, a systematic review in the Arab world had also found a wide range of prevalence estimates for the three MBI subscales across studies on medical professionals ranging between (20.0–81.0%) for high EE, (9.2–80.0%) high DP, and (13.3– 85.8%) for low PA (43). Although there are variations in the reported prevalence of burnout among residents across different studies, attributed to the different factors such as country of residence, specialty, ongoing collective stressful events, and measurement tool used, our findings suggest that burnout may be more widespread than previously thought, potentially worsening as crises become chronic and as they accumulate (44–47).

In line with recent findings, our study further shows that frequent interaction with infected patients is a risk factor for burnout, EE dimension in specific. The impact of interacting with COVID-19 patients on levels of burnout and mental health outcomes of residents has been systematically documented in the literature (48). This was indeed one of the highlights of the General Medical Council national survey in 2021 reporting that one third of trainees said they felt burnt out to a high or very high degree because of their work, compared to around a quarter in previous years (49), with 43% of respondents finding their work emotionally exhausting to a high or very high degree(49). A cross-sectional study in Japan additionally found a positive correlation between burnout levels and the actual number of COVID-19 patients the residents handled (50). Keeping in mind that the impact of COVID-19 pandemic on burnout persists for years after the initial interaction, as documented by a national survey involving radiology residents(51) our findings warrant adequate follow-up on the mental health of impacted residents.

Amidst the Lebanese context, trainees faced the dramatic interplay of many crises including the COVID-19 pandemic, the Beirut blast, and an unprecedented economic crisis. Those who felt burdened due to their involvement in educational activities and those who were physically affected by the blast surprisingly had a higher sense of personal achievement. A possible explanation could be that overcoming physical challenges or surviving a traumatic event would foster a sense of resilience and accomplishment. Facing significant adversities and coping with harsh situations might enhance one’s perception of his/her own capabilities and achievements, leading to a stronger sense of PA and subsequently buffer burnout (52). In our study, we found significant demographic factors associated with increased risk for burnout such as being a male, living alone or with a partner, and having a low income. A similar study in Lebanon had recently documented the deleterious impact of ramping economic worries on individuals’ wellbeing amidst the COVID-19 pandemic; worsened by the country’s fragile structural financial status (53). The study further reported that men had increased financial anxiety compared to women as they are traditionally primarily tasked with providing for their families in Lebanon (53). A study on medical staff in Shanghai reached similar results regarding living arrangements (54). Specifically, being single or married without children (equivalent to living with a partner) was associated with higher levels of DP (54). The financial impact on the burnout of healthcare workers is also a consistent finding across several studies (55–58). While having a low income was a risk factor for burnout in our study, rewarding and fair payment was shown to protect against physician’s burnout (59).

Positive screening for anxiety and depression symptoms was also noted in most of the study participants. This is in line with concurrent studies in the general population indicating similar mental distress rates in young adults in Lebanon during the same period (60,61). These high prevalence rates of mental distress in our sample could primarily reflect the mental cost of adjusting to accumulating crises and should be monitored longitudinally, especially in medical residents. We further intriguingly found that elevated symptoms of mental distress correlated with decreased DP but with increased occurrences of EE and low PA. These findings first and foremost highlight the complex relationship between mental distress and aspects of burnout in the medical residents particularly. As such, it could be that residents experiencing symptoms of pervasive anxiety and depression might overly focus on their work to get distracted from their own struggles, increasing connection to their patients and thus decreasing cynical DP while incurring a cost in terms of emotional weariness and depletion and a decreased overall satisfaction with one’s achievements. In this regard, anxiety and depression could be viewed as making individuals more aware of their own emotions and struggles, and would as such reduce the likelihood of becoming distant and impersonal from patients(62–64). Taken altogether, these findings reflect a sizable need to implement institutional collective strategies to alleviate personal struggles.

### Limitations and Strengths

The cross-sectional study design is a limitation of this study as it does not allow the evaluation of causality or analyze a possible temporal relationship between outcomes. Moreover, exposure to COVID-19 patients and to the blast were assessed using questions that are subjective rather than using validated scales. Amid this, self-reflection bias will also be a limitation. The third limitation is the possibility of recall bias, as questionnaires were filled at a time distant from the exposure event. The participants were recruited through social media platforms which doesn’t allow proper stratification of the sample to ensure better generalizability. A larger sample would have provided more robust associations among the examined variables. The sample size was limited due to challenges concerning the response rate which is expected in such studies due to the ongoing stigma on mental health in Lebanon and concerns about confidentiality. Despite the above limitations, this study has several strengths. To our knowledge, this is one of the few studies on medical residents that draws upon the SDT framework and determines the interplay of self-determination and the various dimensions of burnout. It presents a comprehensive examination of burnout prevalence among medical residents in Lebanon, particularly amidst the unique challenges posed by multiple concurrent crises using validated and well-established tools in the literature. Unlike previous research that has predominantly focused on burnout within conventional stressors, our study delves into the multifaceted impact of these crises on the wellbeing of residents. By exploring these factors within the Lebanese context, we aimed to provide a nuanced understanding of the mechanisms underlying burnout among medical residents, what protects against it and its implications for mental health outcomes.

### Conclusions and Implications for Future Research

The study sheds light on the important relationship of self-determination with the three dimensions of burnout. Self-determination seems to be associated with more DP but with less EE and low PA, the latter being associated with more advanced mental distress, anxiety and depression symptomatology. The higher prevalence of burnout among the medical-resident population in Lebanon as compared to others elsewhere confirms the deleterious effects external stressors have on burnout. Better understanding of burnout among medical residents, a vulnerable but essential workforce on the medical front, would lead to better outcomes and better mental wellbeing of this population. This is in the best interests not only of patients, but the healthcare system and society. Further research should be done to better understand this multifactorial phenomenon and application of tools that enhance self-determination and improve residents’ resilience against the heightened levels of stress inherent in residency.

## Data Availability

The datasets used and/or analyses during the current study are available from the corresponding author on reasonable request.

## Declarations

### Ethics approval and consent to participate

All participants approved the informed consent given by the Institutional Review Board at LAU (LAU.SOM.RC1.30/Dec/2020) before filling the survey. The informed consent for participation in the study was obtained in electronically. All participants first provided a written consent by approving to participate and then were required to fill the anonymized online survey.

### Authors’ contributions

R.C, M.EM and E.T designed the study. E.T helped with data collection. H.M and R.C wrote the main manuscript and C.H and P.S edited it. and prepared the analyses and corresponding tables and figures. All authors reviewed the manuscript.

## Acknowledgements

The authors wish to acknowledge all those who made the project possible.

